# Anterior Spinal Artery Syndrome due to Intervertebral Disc Herniation

**DOI:** 10.1101/2021.03.18.21253916

**Authors:** Asraful Islam, Mohammad D. Hossain, Abu Bakar Siddik, Tyfur Rahman, Ashraful Alam, Md.Manjurul Islam Shourav, Nahian Afrida, Sajedur Rahman, Masum Rahman

**Author notes:** Conflict of interest: All authors declare no conflict of interest. Disclosures: The authors have no financial conflicts of interest to declare. Human/Animal Rights: This article does not contain any studies with human or animal subjects performed by any of the authors.

## Abstract

**Objective:** Anterior spinal artery syndrome (ASAS) has been rarely reported as a complication of intervertebral disc herniation (IVDH). Precipitation factors, presentation, evaluation, treatment strategy, and degrees of recovery have not yet been well documented.

**Methods:** Systematic review was conducted according to PRISMA guidelines to review and summarize for the qualitative synthesis of the data from reported cases of anterior spinal artery syndrome due to intervertebral disc herniation from 1980 to February 2021.

**Results:** A total of 12 cases were reviewed, the median age was 48.5 years. Motor weakness with or without pain was the most frequent presenting symptom accompanying bowel or bladder incontinence (25%) or diminished pain and temperature sensation with spared dorsal column sensation. 40% of conservatively treated patients had complete recovery without any residual deficit. Whereas all patients who managed surgically regained fully functional status with shorter recovery intervals.

**Conclusion:** Abrupt onset of motor weakness is a potential warning symptom of spinal cord infarction, rarely attributed to ASA compression by a herniated disc. Moreover, an accompanying diminished pain and temperature sensation with spared dorsal column sensation is further intimation. Reestablishment of blood flow may bear a favorable outcome.

## 1. Introduction

The spinal cord receives its blood supply by three longitudinal spinal arteries along its entire length, one anterior and two posterior spinal arteries. Segmental arteries further reinforce this circulation via radicular arteries below the cervical cord level [1].T4-T9 segments of the anterior thoracic cord are regarded as a watershed area, leaving it more vulnerable to ischemia. Of concern, the artery of Adamkiewicz, quite often the lowest radicular feeder, emerges from the T9-L1, reaches the dural sac, ascends one to three segments to join the anterior spinal artery[2].Acute ischemic cord injury, either from occlusive or nonocclusive hypoperfusion of the anterior spinal artery (ASA), is referred to as the anterior spinal artery syndrome or ASAS[3]. Pathologies affecting the anterior spinal artery or ASA commonly include arteriovenous malformation, vasculitis, iatrogenic causes[4], [5]. Rarer causes include hyperextension positioning[6], fibrocartilaginous embolism, thoracic disk herniation, and cervical spondylosis[7]. Anterior spinal artery syndrome (ASAS) secondary to disk herniation yields only a few cases reported so far.

In most instances, herniated intervertebral disc leads to spinal cord pathology by direct compression of the cord [8]. This is manifested by sudden-onset dorsalgia, dermatomal sensory deficits, and myelopathy, which are most often slowly progressive over months to years. It has reported cord ischemia presenting as either ASAS or Brown-Sequard syndrome secondary to disc herniation. The latter is thought to be attributed to a spinal branch of a segmental artery compression. In contrast, ASAS is most likely triggered by the complete or incomplete occlusion of the ASA or its feeder artery attributed to the disc herniation [5].

ASA syndrome in the setting of intervertebral disc herniation is a rarely reported form of cord ischemia. While the overall clinical outcome of ASAS is poor, degrees of recovery related to a disc herniation is yet to explore. The management options for ASAs from disc herniation are also ambivalent. It is unclear if any benefit of steroid and heparin treatment, timing, and role of decompressive surgery is also inconclusive. Herein, we conducted a comprehensive review of existing literature to answer the following questions: 1) What are the precipitating factors? 2) What is the distinct manner of presentation? 3) What are the characteristic imaging findings 4) What regions of the spine frequently involve? 5) What treatment strategies are administered? 6) What are the degrees of recovery?

### 2. Methodology

The research protocol was decided at the start of the project, which specified goals, search design, inclusion/exclusion criteria, data extraction, outcomes, and analysis techniques.

### 2.1. Study design

A systematic literature review was conducted under the PRISMA (Preferred Reporting Items for Systematic Reviews and Meta-Analyses) guidelines to identify studies anterior spinal artery secondary to due to intervertebral disc herniation. Formulated PICO question for the review described, Population: patient clinical and imaging evidence of anterior spinal artery syndrome by intervertebral disc herniation; Intervention: Surgical decompression or conservative management, Compare: no comparator; Outcome: long term outcome, residual neuro deficit, rate of recovery, speed of recovery.

### 2.2 Search strategy

Databases used for comprehensive literature search include Ovid MEDLINE(R) and Epub Ahead of Print, In-Process and Other Non-Indexed Citations, and Daily, Ovid EMBASE, Ovid Cochrane Central Register of Controlled Trials, Ovid Cochrane Database of Systematic Reviews, and Scopus. The search design was planned and carried out by a medical reference librarian with feedback from the investigators. Search for anterior spinal artery syndrome due to intervertebral disk herniation was completed by using the following terms: “intervertebral disc,” “spondylosis,” “Spinal cord infarct,” “anterior Spinal Artery,” “anterior spinal artery syndrome,” “ASA,” “ASAS,” “Spinal cord ischemia,” “Spinal Artery dissection,” “Spinal Artery occlusion,” “Spinal Artery compression,” “ASAS” along with “AND” and “OR” operators and published in the English language.

### 2.3. Eligibility criteria

We included case reports and other studies in which anterior cord ischemia was evident most likely due to ASA occlusion by herniated intervertebral disc were included. Due to a very scant number of cases, we also collected data from some review articles that reported several non-english literature cases, and those were not included in our primary search results. No restrictions were applied to age, gender, ethnicity, and geographical location. The reference list of included papers was examined for other pertinent studies not identified by our initial search terms. Studies were excluded if in animal models or without patient data, not fulfilling the study population criteria, conference abstracts only, review article or meta-analysis in the absence of new patient data, or studies without full text. The literature search strategy and paper inclusion were performed by two authors independently

### 2.4. Study selection process

Duplicate studies have been removed from all citations received by extensive literature searches. Titles and abstracts have been selected for inclusion by pairs of researchers. The same pairs of reviewers recorded abstracts, also screened and retrieved full text. Conflicts have been resolved by the third independent reviewer.

### 2.5. Data collection

Two independent authors extracted data from finally selected studies in a pre-piloted electronic database. Due to a very scant number of cases, we also collected data from some review articles that reported several non-English literatures cases and were not included in our primary search results. The literature search strategy and paper inclusion were performed by two authors independently. Patient demographic data, possible precipitating factors, duration of neuro deficits, initial presentation, the pattern of motor and sensory deficit, physical findings, imaging data, treatment approaches, outcome, follow-up data were collected for qualitative synthesis. To reduce the chances of bias, no attention was paid to the author, journal, or country of origin, and the outcome data were recorded according to the patient perspective.

## 3. Results

A total 12 cases from twelve papers were listed for analysis after a systematic literature search was performed based on the defined inclusion criteria [5], [7], [9]– [18]. The search results are summarized in a PRISMA diagram. The studies that were included were then summarized in tables. The cases included had clinical and MRI features of ASAS and intervertebral disc herniation [table 1] with a median age of 48.5 years (Range 31 years - 82 years) and male to female ratio was 1:1. All of the patients presented without any precipitating trauma; however, straining and abrupt blood pressure reduction by beta-blocker was thought to be an initiator in two cases. Duration of symptoms was reported in 8 cases; rest reported as acute, on average 31.25 hours (range: 1-168 hours). To make the cohort homogenous, we have categorized acute when presented within 24 hours. To that, 11 (91.66%) patients presented acutely, and the shortest duration was 1 hour.

**Table 1:** Patient demographics and presentation

| No. | Author | Patients | Duration of neurodeficits (hours) | Motor disturbance | Sensory disturbance | Bowel or bladder disturbance | Clinical diagnosis of ASAS |
| --- | --- | --- | --- | --- | --- | --- | --- |
| 1 | Santillan A | 54, F | 24 | Paraparesis | Bilateral Hoffman's sign, diminished pinprick sensation below T3, saddle anesthesia | Yes | Yes |
| 2 | Peng T | 31, M | 24 | Quadriparesis | Loss of light touch and pin prick sensation at the level of C5 | No | Yes |
| 3 | Acker G | 72, F | 5 | Quadriparesis | Disturbed thermal perception. The gait was slightly ataxic. | No | Yes |
| 4 | Reynolds JM | 36, F | 12 | Paraparesis | Crude touch, pinprick, and temperature were absent bilaterally from T8 and below | No | Yes |
| 5 | Li Y | 82, M | 4 | Quadriplegia | Sensations were hyperaesthetic in the left C4 and C5 dermatomes | No | Yes |
| 6 | Guest JD | 38, F | 12 | Paraplegia | Decrease T4 sensory level to pinprick and temperature | Yes | Yes |
| 7 | La Torre E | 43, M | 168 | Quadriparesis | Touch, pain, temperature sensation decreases in right hand. | Yes | Yes |
| 8 | Gregory S Schneider | 48, M | 1 | Quadriparesis | Loss of light touch and pin prick sensation at the level of C6. | No | Yes |
| 9 | Errea | 49, M | N/A | Quadriparesis | Lancinating interscapular pain | N/A | Yes |
| 10 | Odaka | 54, F | N/A | Quadriparesis | Angina | N/A | Yes |
| 11 | Arai | 40, F | N/A | Quadriparesis | NA | N/A | Yes |
| 12 | Yano | 78, M | N/A | Paraplegia | NA | N/A | Yes |

**Table 2:** Evaluation, management and clinical outcome

| No. | Author | Patients | Cord ischemia on imaging | Level of disc herniation | Evidence of ASA occlusion | Steroid therapy | Surgical decompression | Physiotherapy | Recovery | Duration of recovery (Days) |
| --- | --- | --- | --- | --- | --- | --- | --- | --- | --- | --- |
| 1 | Santillan A | 54, F | Yes | T8/T9 | Yes | No | Yes | No | Complete | 5 |
| 2 | Peng T | 31, M | Yes | C3,4 and C4,5 | Yes | Yes | Yes | No | Complete | 7 |
| 3 | Acker G | 72, F | Yes | C5/6 | NA | 1 (8mg TID) | Yes | No | Complete | 63 |
| 4 | Reynolds JM | 36, F | Yes | T7–T8 | No | 1 (1g daily for 5 days) | No | Yes | Partial | 90 |
| 5 | li Y | 82, M | Yes | C3/4 to C6/7 | No | Yes | No | Yes | No Recovery |  |
| 6 | Guest JD | 38, F | Yes | T8/9 | Yes | Yes | No | No | Complete | 10 |
| 7 | La Torre E | 43, M | No | C5-C6 | No | No | Yes | No | Complete | 18 |
| 8 | Gregory S Schneider | 48, M | No | C6–7 | No | Yes | No | No | No Recovery | NA |
| 9 | Errea | 49, M | Yes | C6,7 | N/A | N/A | Yes | N/A | N/A | N/A |
| 10 | Odaka | 54, F | Yes | C5-C7 | N/A | N/A | Yes | N/A | N/A | N/A |
| 11 | Arai | 40, F | Yes | C4,5 | Yes | N/A | Yes | N/A | N/A | N/A |
| 12 | Yano | 78, M | Yes | T8,9 | No | N/A | No | N/A | N/A | N/A |

NA = Not available

Keeping consistency with ASA syndrome, all patients had motor weakness, 4 (33%) presented with paraparesis and 8 with quadriparesis (67%). Sensory disturbances were primarily diminished pain and temperature sensation with spared dorsal column sensation, characteristic of ASAS. 25% of the study population (n=3) had bowel or bladder incontinence in their presentation. All three have returned to normal functioning of bowel and bladder on follow up.

10 out of 12 (83%) study population had imaging findings consistent for spinal cord ischemia of ASA distribution and all of the patients had concomitant disc herniation. In fact, angiographic evidence ASA occlusion is the gold standard for diagnosis; data was available in 9 cases (83 %) where complete occlusion wasn’t reported in 1 instance.

Spinal cord infarction is typically managed conservatively. Corticosteroid therapy has not been established in acute spinal cord ischemia [19]; however, it might be justified where accompanying compression is evident. Regardless, revascularization is the treatment of choice when present within a salvageable window. Surgical decompression, therefore, should be offered for ASA occlusion by a herniated disc. In our study, 58% (n=7) underwent surgical decompression, and 2 patients (16%) received adjuvant steroid therapy, rest managed conservatively with steroid only. Data for steroid therapy wasn’t available for 4 of the patients.

Complete recovery without any residual deficit was achieved in 100% of cases managed surgically and 40% managed without surgical decompression. 40% of conservatively treated cases demonstrated partial recovery, and the remaining 20% showed no neurological improvement at all. The recovery period for the surgery cohort was ranging from 5 days to 63 days and 10 to 90 days in the conservative cohort. The average followed-up length for the reported cases 11.6 months.

## 4. Discussion

Spinal cord infarction is reasonably uncommon and represents 0.3–1 % of all CNS infarctions. However, recent studies show 14 to 18% of transverse myelitis cases are secondary to ischemic myelopathy, implicating the possibilities of underdiagnosis.[20],[21] Among all, ASAS is the most common form and reported up to 87.2% of the cases. Reportedly, cord ischemia presents with a wide age distribution ranging from the first to the tenth decade of life, with a median age between 50 and 70years. Although gender preference is inconclusive for ASAS, elderly females demonstrated the worst prognosis[20], [21]. In our analysis, the median age was 48.5years with a range of 31 years to 82 years with equal sex distribution.

### 4.1. Pathogenesis

Anterior spinal artery (ASA) arises from the vertebral arteries at the level of the medulla. It runs down rostrally along the anterior median sulcus, supplies the anterior portion of the spinal cord. Pathophysiologically, generalized hypoperfusion or localized vascular interruption accounts for spinal cord infarction. Common contributors for ASAS include thoracoabdominal aortic surgery, spinal surgery, aortic dissection or aneurysm, atherosclerosis, vasculitis, arteriovenous malformation (AVM), embolic causes, and hypercoagulability. Symptomatic ASAS due to a herniated intervertebral disc has been rarely reported and thought to have occurred in the setting of direct compression to ASA or its feeding artery [3],[22].

Ischemic change is the critical contributor to the pathogenesis for spondylotic myelopathy is well established [23]. Morphological changes imply ischemic insult include necrosis, edema, myelomalacia, gliosis, and inflammation [24]. Spondylotic myelopathy, therefore, stems from either ischemia due to higher interstitial pressure by direct cord compression or arterial occlusion by a herniated disc. In a study on 14 patients with cervical spondylotic myelopathy, no ASA occlusion was evident by CT angiography, even with 80% sagittal diameter compression and T2 hyperintensity [25]. Thus, compression of the spinal cord on its anterior surface is more probable than ASA. Nevertheless, more patient groups are needed to identify and study. Considering true arterial occlusion, a typical posterolaterally displaced disc is more likely to compress the radicular artery. However, direct compression to ASA has also been reported, should be considered as more severe than occluding its feeding branch [5], [11], [12], [14], [15]. Therefore, a probability of ASA interruption should be considered in a spondylotic patient with ventral cord syndrome.

In our study, disc herniation was demonstrated in a diverse range of vertebral levels. The cervical region was most frequently involved in 66% of cases, and the remaining 34% were in the lower thoracic(T7-T12) region. The degree of motor and sensory deficit was consistent with the level of injury. Patients who had disc-herniation in cervical vertebrae manifested quadriparesis, while paraparesis for the thoracic region.

### 4.2. Clinical presentation

ASAS presents as an abrupt onset of painful myelopathy with bilateral loss of motor functions, pain, and temperature sensation. Symptoms typically appear with acute back or neck pain, referring to the level of injury and neurological impairment progress within 2-24 hours. Depending on the level of infarction, motor impairment commonly demonstrates as paraplegia or quadriplegia. Due to ventral horn injury or spinal shock, flaccid motor paralysis with decreased deep tendon reflexes is typical at the beginning. However, over time, patients progress to upper motor signs such as hyperactive reflexes and spasticity. Sensory disturbances typically include loss of pain and temperature sensation, preserved dorsal column sensation, vibratory sense, and proprioception. Patients may also experience autonomic dysfunction complicated by bowel and bladder dysfunction as well as hypotension [4], [10], [26].

In our analysis, all of the cases present with motor weakness, 67% with quadriparesis and 33% paraparesis; among them, 60% had pain at presentation. Although not presented as a symptom, sensory disturbances were evident in neurological examinations. This includes loss of perception of pain and temperature with preserved dorsal column sensation, vibratory sense, and proprioception unique to ASAS. 25% of our cases also had bowel or bladder disturbances at the presentation.

### 4.3. Imaging

The diagnosis of ASAS was based mostly on clinical presentation and imaging study. MRI is the most sensitive and reliable imaging modality to detect spinal cord infarction [3]. Within a few hours of the onset of symptoms, it can detect signal changes and swelling related to cord ischemia and also demonstrate other pathologies, such as extradural compression, intramedullary tumors, AVM, and multiple sclerosis plaques. The typical finding is T2 hyperintensity resembling the “owl-eye” or “snake-eye” pattern in axial planes and “pencil-like” T2 hyperintensity over multiple segments in the vertical plane. In contrast to conventional sequences, DWI can detect cord ischemia within a few minutes following symptom onset [10], [22], [27]. Cervical spondylosis may occupy over 80% sagittal diameter of the spinal canal and exhibit T2 hyperintensity on MRI. Therefore, ASAS in the setting of spondylosis should not solely be based on clinical findings preferably on ASA occlusion [25]. In our analysis, angiographic evidence of ASAS was not reported in all cases. However, primary authors of reported cases were concluded ASAS from a herniated disc by clinical and imaging evidence of concurrent spinal ischemia and disc herniation. We recommend angiograms in every case of ASAS to detect any external compression to justify the decision for surgical decompression.

### 4.4. Management and outcome

The exact cause of ventral cord syndrome directs the management plan. Acute bulging of an intervertebral disk may cause ventral cord dysfunction with or without ASA compression. However, ASA occlusion explains ischemic cord injury rather than direct compression. Systemic steroids have no beneficial effects on ischemic spinal cord injury. Early intervention to prevent pulmonary, hemodynamic, and thromboembolic complications is critical in acute management. Long-term neurological outcome solely depends on the reversing of the ASA decompression and re-establishment of blood flow. Therefore, the timing between the occlusion and intervention is the most critical factor for long-term prognosis. Early initiation of physiotherapy and extensive rehabilitation training has a potential role in regaining neurologic function, but these interventions are not superior to urgent decompression surgery. Regardless, the approaches of imaging and interventions must be explored much more diligently to ensure patient benefit, reduce the degree of morbidity, impairment, and an increase in psychological well-being.

Seven patients underwent surgical treatment to decompress the anterior spinal artery; all of them demonstrated complete recovery, and the mean duration of the recovery period was 23.25 days (5-63days). In contrast, full recovery occurred only in 40% of cases managed conservatively with the longest recovery length, 90days. Hypothetically revascularization should have a better prognostic outcome. However, the interval between onset and intervention, use of steroids, other vascular or spinal comorbidities may have the potential influence yet need to be studied.

A diverse array of recovery is possible following infarction of the spinal cord. A study on spinal cord stroke demonstrated that recovery rate of walking power was 41% for those using a wheelchair at discharge and rate of removal of catheter was 33% for those with a long-term catheter in situ [28].

## 5. Limitations

This study has several methodological limitations, such as insufficient sample size, selection criteria, inadequate follow-up data, and comparator. Population size is too small for statistical measurement and conclusion. Besides this, lack of angiogram data may include at least a few unrelated patients, where ASA occlusion might not cause spinal cord infarction. Due to lack of sufficient data, we couldn’t identify the risk factors or precipitating factors for this condition; thus, it still remains a question when to suspect. It’s an observational study on existing reported cases; thus, cases were not randomly distributed for different treatment approaches with having a control group. Hence, better outcome in specific population is still in hypothesis until reproduced in further clinical studies.

## 6. Conclusion

Spinal cord infarction due to herniated intervertebral disc induced ASAS is possible and maybe under-evaluated, also under-reported. A thorough evaluation of patients’ symptoms, as well as a neurological examination to detect distinguishing signs, would help in the early diagnosis of this unusual disease. When suspicion arises, all patients should have an angiogram to identify ASA occlusion to identify the potential candidate for surgical decompression.

## Data Availability

All data are available in figures and tables.

## Acknowledgments

We want to acknowledge and thank Larry J. Prokop, Outreach Librarian, Mayo Clinic Libraries, and CMSR (center for medical study and research) foundation.

---

**Figure 1.**
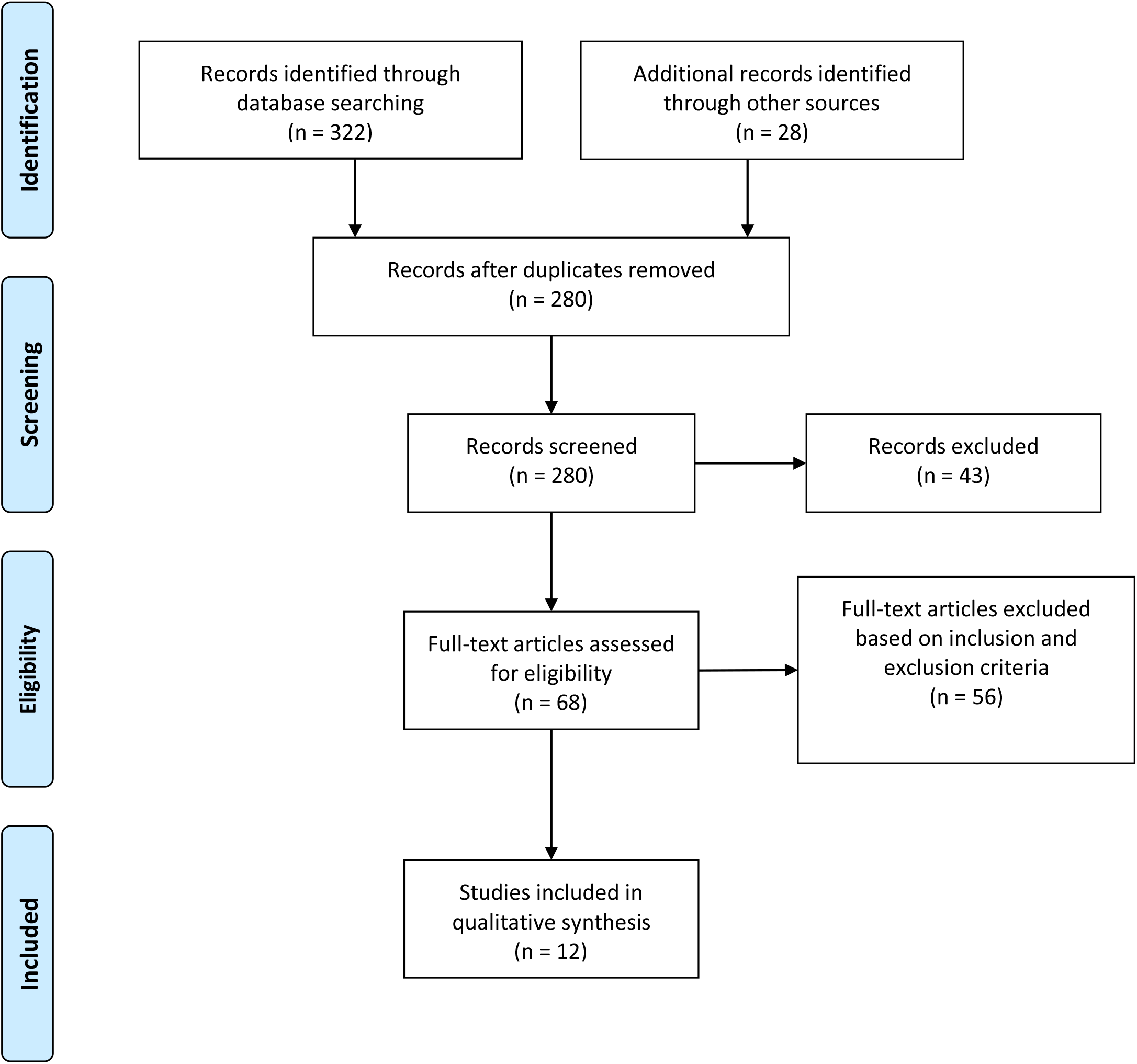
PRISMA flow chart used for study selection relevant to anterior spinal artery syndrome secondary to intervertebral disc herniation.

**Figure 2:**
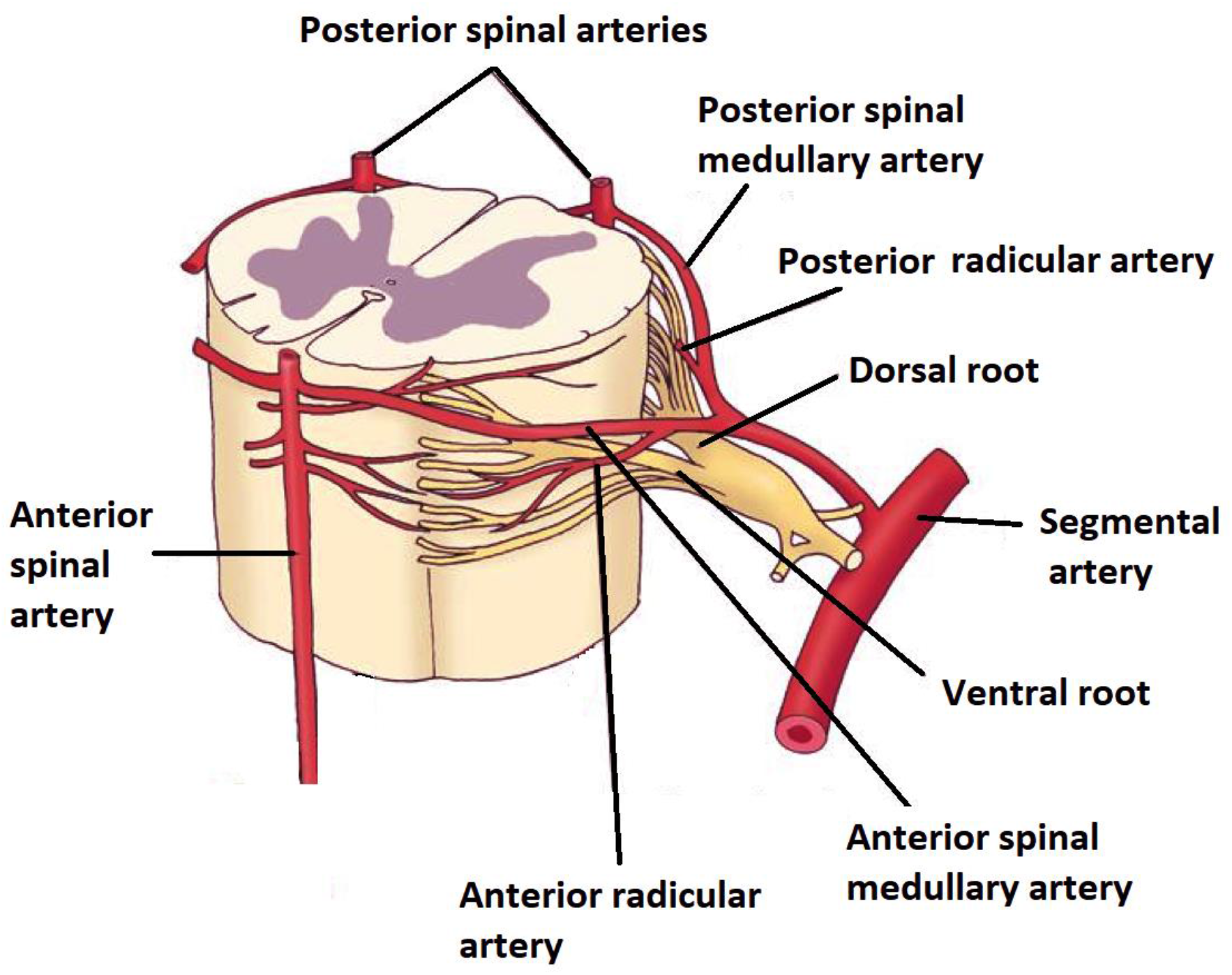
Diagrammatic representation of segmental spinal cord blood supply. Adopted and modified from lippincott williams and wilkins. 2006.

**Figure 3.**
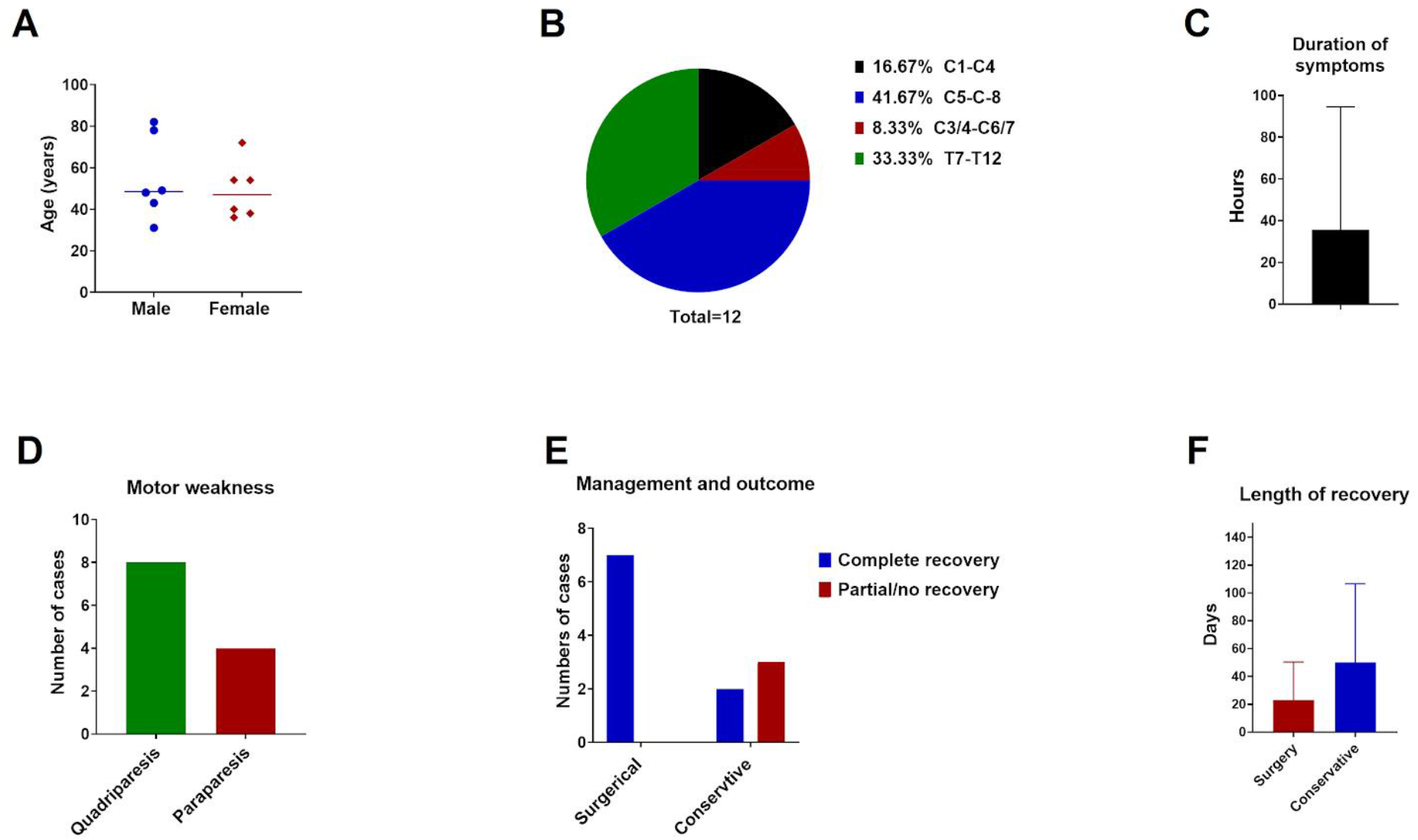
Patient demographic and clinical data. A: Age and sex distribution of ASAS due to IVDH, showing equal sex distribution and a median age of 48.5 years. B: Frequency distribution of level of lesion. C: Symptom durations of all presented cases. D: Pattern of motor weakness reported. E-F: Management plan and related outcome data.

**Figure 4.**
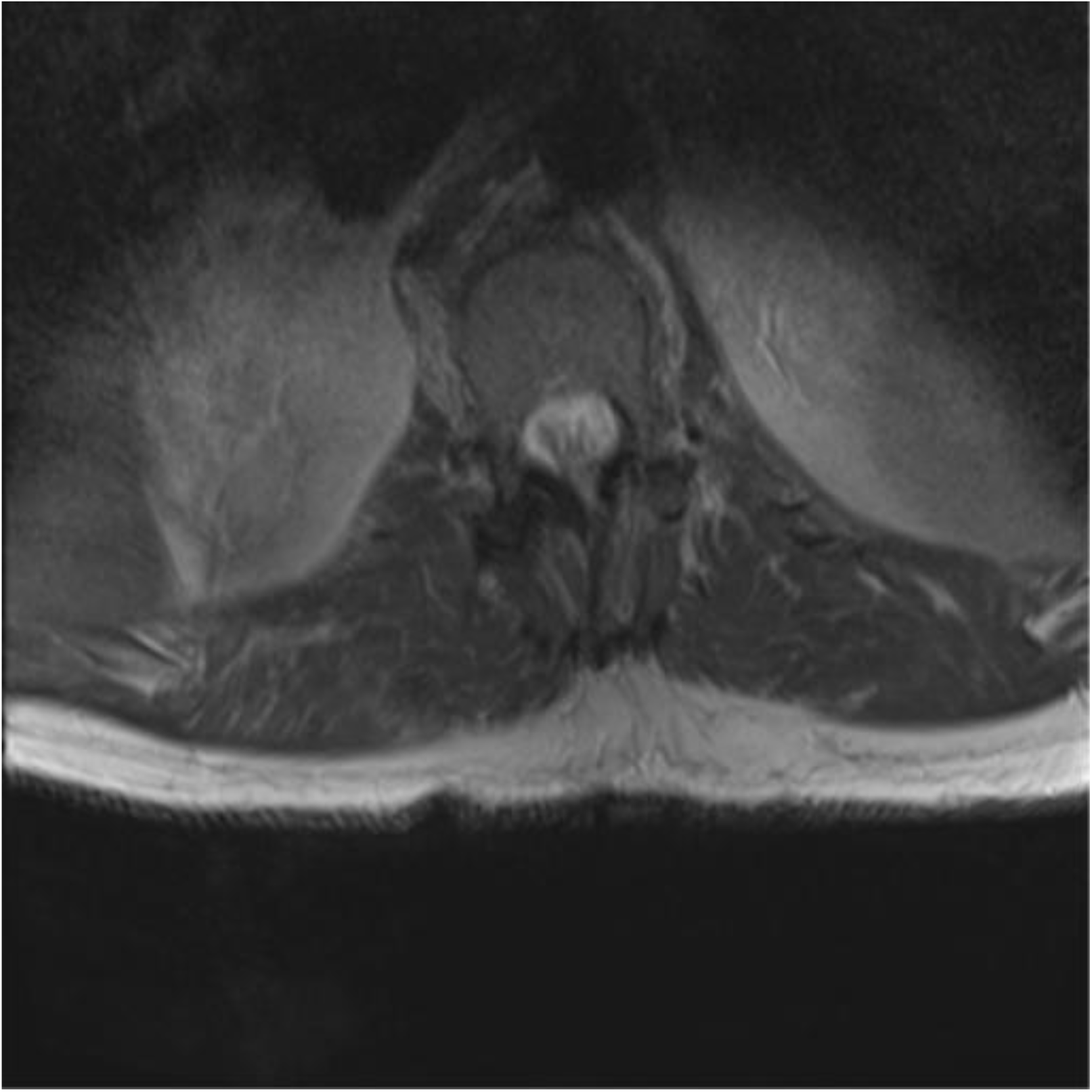
High signal intensity on T2 weighted image, demonstrating anterior cord infarction. Rest of the cord has normal signal intensity and caliber with sizeable canal and intervertebral foramina. Patient presented with traumatic thoracic aortic injury treated TEVAR and had clinical features of anterior cord syndrome. Case courtesy of Dr Bruno Di Muzio, Source: radiopaedia.org/cases/57072.

